# Luzon, Visayas, and Mindanao (LuzViMin) During COVID-19: Comparative Analysis on Epidemiology, Pandemic Mobility and Evolutionary Dynamics in the Three Major Islands in the Philippines

**DOI:** 10.1101/2025.08.07.25333229

**Authors:** Joaquin Antonio B. Limbo, Abegail V. Madrigal, Gorgeous Ivana C. Lim, Luisa Marie S. Jingco, John Kim D. Aligato, Richard C. Cipriano, Kristine Pearle E. Cabildo, Elaine Mae C. Allosa, Addie Franchesca D. Manrique, Daniel Jericho H. Dungo, Albert Derrick R. Recio, Joe Anthony H. Manzano, Nicanor Austriaco OP

## Abstract

The COVID-19 pandemic exposed the vulnerabilities of developing nations, particularly the Philippines, where geographic fragmentation and socio-demographic diversity posed unique challenges to disease control. As an archipelagic country composed of Luzon, Visayas, and Mindanao, the Philippines presents a distinct opportunity to examine how geographic, political, and cultural factors shape pandemic dynamics. While previous studies have focused on SARS-CoV-2 genomics, few have integrated epidemiological trends, mobility patterns, and viral evolution across the islands. This study addresses that gap by analyzing data from January 2020 to June 2022 to identify island-specific variations in COVID-19 transmission and response. Epidemiological records, mobility data, and SARS-CoV-2 genome sequences were processed using Python and R, with data completion *via* forward-filling and linear interpolation. Phylogenetic analyses were performed using Nextstrain. Results showed that early lockdowns reduced cases and deaths across all islands, with Luzon experiencing the most sustained improvements. Visayas and Mindanao showed higher mobility throughout, which may have contributed to delayed case reductions and greater mortality during the Delta wave. Luzon also saw earlier and sharper declines in severe cases and fatalities, possibly due to more efficient vaccine rollout. While overall viral evolution was similar across regions, minor genomic variations were noted in Visayas and Mindanao. This study offers the first comparative assessment of epidemiological, mobility, and genomic patterns across the three major Philippine islands. Findings emphasize the need to tailor public health strategies to regional contexts, supporting more responsive and resilient pandemic preparedness across geographically diverse nations.

## INTRODUCTION

The COVID-19 pandemic is one of the most significant global health crises in recent history, revealing critical weaknesses in healthcare systems, economies, and societal infrastructures across the world (World Health Organization 2020). Developing countries, in particular, experienced disproportionate impacts due to limited healthcare resources, high population density, and long-standing socio-economic inequalities (World Bank 2020). In this context, the Philippines presents a unique case. As an archipelagic nation composed of three major island groups Luzon, Visayas, and Mindanao, the country experienced the pandemic not as a single uniform event, but as a set of regionally distinct outbreaks shaped by geography, governance, and population behavior (Mata et al. 2024; Otero et al. 2025).

Each island group possesses its own geographical, cultural, political, and socio-demographic features that likely influenced the transmission dynamics of SARS-CoV-2 and the effectiveness of local pandemic responses (Dimaano et al. 2024). While genomic surveillance and evolutionary studies of SARS-CoV-2 in the Philippines have been conducted (Tablizo et al. 2021; Dimaano et al. 2024; Velasco et al. 2020), research that integrates regional epidemiological data with human mobility trends and viral genomic evolution remains limited. This lack of comparative, island-specific analysis poses a significant challenge to evidence-based policymaking, particularly for future public health emergencies.

Previous studies, such as Mata et al. in 2024, Dimaano et al. in 2024, and Li et al. in 2022, have primarily focused on national-level trends, which often overlook the nuances of regional variation. For instance, Luzon, the country’s most populous and economically developed region, was able to implement early and strict mobility restrictions that may have contributed to a faster reduction in COVID-19 cases and mortality rates. On the other hand, Visayas and Mindanao reported higher mobility and slower declines in cases and deaths, which may reflect disparities in access to healthcare, vaccination rollout, and local enforcement of public health measures (Dimaano et al., 2024). These regional differences suggest that a more granular analysis is needed to better understand how local contexts shaped the pandemic trajectory.

Additionally, while phylogenetic studies have provided insights into the broader evolutionary landscape of the virus within the Philippines (Tablizo et al. 2021), little is known about how these evolutionary dynamics varied across Luzon, Visayas, and Mindanao. This information is essential in identifying regional transmission clusters, mutation patterns, and potential hotspots for variant emergence. The interaction between mobility behavior, epidemiological trends, and viral evolution remains underexplored, especially in geographically complex countries like the Philippines.

This study aims to address these gaps by conducting a comparative analysis of pandemic-related epidemiological patterns, mobility trends, and evolutionary dynamics of SARS-CoV-2 across Luzon, Visayas, and Mindanao. Using data collected from January 2020 to June 2022, we integrate case counts, mobility indices, and viral genome sequences to explore how the pandemic unfolded in each of the three island groups. Specifically, we examine (1) regional variations in COVID-19 case and mortality trends, (2) the relationship between mobility behavior and transmission dynamics, and (3) differences in phylogenetic patterns across regions. By offering the first integrated, region-focused analysis of the COVID-19 pandemic in the Philippines, this study contributes to a deeper understanding of how geographic and socio-demographic differences influence pandemic outcomes. The findings underscore the importance of localized and data-driven public health strategies, both for ongoing response and for future pandemic preparedness in the Philippines and other archipelagic or regionally diverse nations.

## MATERIALS AND METHODS

### Data collection

This study utilized publicly available data from the Department of Health (DOH) of the Philippines and open-access news publications. The DOH data included daily reported COVID-19 cases, deaths, and recoveries across major administrative regions, serving as the foundation for epidemiological trend analysis. The DOH data was filtered to include only collections from January 1, 2020, to January 30, 2022.

To complement this dataset, regional and inter-island mobility data were sourced from Facebook’s Data for Good program which references the methods of Gallego-García et al. (2024). This database provides anonymized, aggregated mobility metrics derived from users who have location history enabled. These data captured temporal movement patterns during the pandemic and were used to examine potential correlations with transmission dynamics and variant introductions.

News media reports were systematically reviewed to corroborate the timing of government interventions, such as community quarantines, travel restrictions, and reopening phases, and to provide contextual insights into public sentiment, policy responses, and behavioral compliance.

### Data curation

Genomic sequences of SARS-CoV-2 were sourced from the GISAID database. The dataset included Variants of Concern (VOCs), namely Alpha, Beta, Gamma, Delta, Omicron, and Theta. Sequences were filtered based on metadata completeness, sequence coverage, and quality scores to ensure high confidence in downstream analyses. Only high-quality sequences with complete location and date information were retained. The genomic sequence data was filtered to only include collections from January 1, 2020, to January 30, 2022. A detailed summary of the curation process, including contributor acknowledgments and inclusion criteria, is provided in the Supplementary Materials.

### Data processing

Epidemiological data from the DOH were downloaded in CSV format and processed using Python. Missing values were imputed using trend-based methods consistent with the temporal structure of the data. Outliers were identified through interquartile range analysis and corrected or flagged for sensitivity checks. Mobility data were aggregated by week and normalized to detect significant deviations in population movement over time. News articles were manually coded and grouped according to type of intervention and geographic scope.

### Analytical tools and techniques

Following data curation, a Python-based pipeline was developed to integrate curated genomic sequences, configuration files, and workflow components necessary for the generation of a genomic surveillance phylogenetic tree using the Nextstrain platform. The pipeline outputs a .json file compatible with Auspice (https://auspice.us), enabling interactive visualization of phylogenetic relationships. Visualization parameters were configured to span from January 1, 2020, to June 30, 2022, with primary filtering based on administrative divisions. Additional regional filters (e.g., Eastern, Western, and Central Visayas) were applied as needed to facilitate region-specific analyses. This interactive visualization allowed for detailed exploration of SARS-CoV-2 variant emergence and spatiotemporal distribution, thereby providing contextual insights into the impact of viral evolution on public health responses. The resulting .json file used in this study is publicly accessible, and customization guidance can be found in the official Nextstrain documentation.

### Visualization and interpretation

Temporal epidemiological trends including confirmed cases, mortality, and population mobility were visualized using Matplotlib, allowing for intuitive temporal comparisons. These were complemented by Nextstrain-based genomic visualizations, which illustrated the phylogenetic relationships and mutation patterns of circulating SARS-CoV-2 lineages in the Philippines. By integrating epidemiological data with viral phylogeny, this approach facilitated a comprehensive understanding of how policy decisions, human behavior, and viral evolution intersected over the course of the pandemic.

## RESULTS AND DISCUSSION

### Spatiotemporal Dynamics of COVID-19 Cases and Mortality Across the Three Major Islands

To examine the temporal and geographic dynamics of COVID-19 in the Philippines, we analyzed daily confirmed cases and deaths from January 2020 to mid-2023 across the three major island groups: Luzon, Visayas, and Mindanao. Using Python’s Matplotlib library, we visualized region-specific epidemic curves (Figure 1A and 1B) and contextualized these trends with key variant-driven waves and public health interventions.

**Figure 1:**
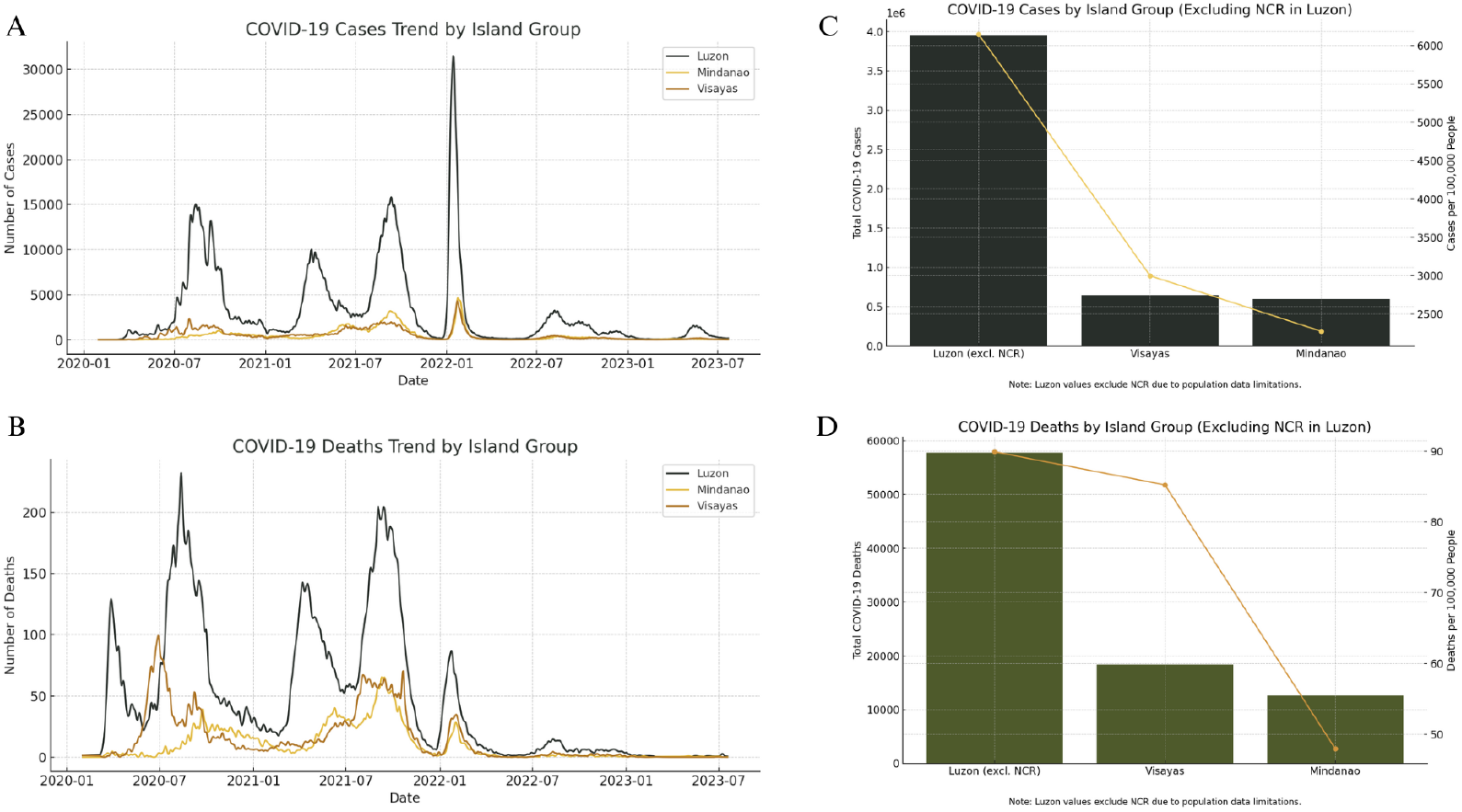
Comparative Graphs of the Reported Cases and Deaths, and Per Capita Cases of COVID-19 in Luzon, Visayas, and Mindanao from 2020-2022. **A**. Daily Reported COVID-19 Cases by Island Group in the Philippines, January 2020 – June 2023. **B**. Daily Reported COVID-19 Deaths by Island Group in the Philippines, January 2020 – June 2023. **C**. Total and Per Capita COVID-19 Cases by Island Group in the Philippines (Excluding NCR in Luzon). **D**. Total and Per Capita COVID-19 Deaths by Island Group in the Philippines (Excluding NCR in Luzon.

The progression of COVID-19 revealed distinct spatial and temporal patterns shaped by factors such as population density, mobility, healthcare infrastructure, and sociopolitical responses. Luzon consistently recorded the highest case counts throughout the pandemic (Figure 1A), largely due to its high population density and the presence of Metro Manila, the country’s primary urban and economic hub. The Omicron-driven surge in January 2022 marked the peak in daily cases, with Luzon reporting over 30,000 infections in a single day. In contrast, Visayas and Mindanao exhibited similar wave patterns during Delta (mid-2021) and Omicron (early 2022), though with markedly lower case burdens. These differences are likely influenced by lower interregional mobility, fewer densely populated urban centers, and varied testing and reporting capacities.

Despite regional disparities in case volume, the synchronized timing of peaks across all three island groups suggests that variant-driven transmission was the dominant factor shaping national epidemic curves. This pattern is consistent with findings from other archipelagic or regionally fragmented countries, such as Indonesia and Japan, where mobility restrictions and regional governance created differential local dynamics, but variants of concern ultimately drove synchronized national waves (Tegally et al. 2022).

Patterns in COVID-19 mortality, however, did not perfectly mirror case incidence, highlighting the complex interplay of viral evolution, healthcare system responses, and public health strategies (Figure 1B). Luzon again reported the highest number of deaths, particularly during the Alpha/Beta and Delta waves. The Delta wave in late 2021 represented the deadliest phase across all regions, likely due to the combination of increased virulence, overwhelmed health systems, and incomplete vaccination coverage at the time. Interestingly, although the Omicron surge led to the highest case numbers, it resulted in significantly fewer deaths. This aligns with global data indicating that the Omicron variant caused milder disease outcomes, especially in populations with high levels of prior immunity from vaccination or past infection (Altarawneh et al. 2022). In the Philippine setting, from mid-2022 onward, both cases and deaths declined steadily in all island groups, suggesting that a combination of increased vaccine coverage, improved clinical management, and population-level immunity helped reduce the public health burden.

The observed regional differences also highlight how sociodemographic factors influenced the epidemic trajectory. For instance, Luzon’s dense urban population and higher international connectivity facilitated faster and wider viral spread, while the more rural, dispersed populations in Visayas and Mindanao likely contributed to slower transmission. Additionally, regional health system capacity and socioeconomic disparities likely affected testing access and the quality of clinical outcomes, echoing global observations that areas with limited healthcare access bore a disproportionate mortality burden (Wachtler et al. 2020).

Mobility patterns further shaped these dynamics. Data from Facebook’s Data for Good mobility reports revealed sharp declines in movement during early lockdowns, with gradual recovery by mid-2021. Regions with lower inbound and outbound mobility, particularly in Mindanao, showed more staggered waves, indicating the role of spatial connectivity in viral dissemination.

### COVID-19 Burden Across Philippine Island Groups Without NCR Influence

To better understand the geographic distribution of COVID-19 burden while minimizing the statistical weight of the National Capital Region (NCR), we performed case and death data analysis with NCR excluded from Luzon (Figures 1C and 1D). This adjustment reveals that even without NCR, Luzon still reported the highest total number of COVID-19 cases, underscoring the significant contributions of other densely populated regions such as CALABARZON and Central Luzon. When adjusted for population, the per capita case rates followed a similar trend with Luzon (excluding NCR) leading the highest number of cases per 100,000 population, followed by Visayas and Mindanao (Figure 1C). These findings suggest that viral transmission in Luzon was not solely concentrated in Metro Manila but extended into semi-urban and rural provinces, likely driven by high economic activity, labor mobility, and extensive inter-provincial connectivity as evidenced by the study of Talabis et al. in 2021. The sustained transmission outside the capital reflects a broader regional vulnerability shaped by both structural and social factors.

A parallel pattern is observed in COVID-19 mortality (Figure 1D), with Luzon (excluding NCR) again exhibiting the highest total and per capita death counts. Although Visayas and Mindanao had lower absolute numbers, the per capita death rate in Visayas surpassed that of Mindanao despite the latter recording more total cases. This discrepancy suggests possible regional differences in healthcare accessibility, hospital capacity, or underlying population health indicators such as comorbidities and age structure. The narrower gap in mortality compared to case incidence also hints at the role of non-pharmaceutical interventions, healthcare quality, and timely access to treatment in mitigating fatal outcomes (Migriño Jr. & Martinez 2025). Collectively, these region-specific patterns reinforce the importance of disaggregated data analysis in understanding pandemic dynamics and inform the need for tailored public health strategies that account for local demographic and healthcare system contexts.

### Mobility Patterns Across Philippine Island Groups

To assess the impact of COVID-19 restrictions on population movement across the Philippine archipelago, we analyzed mobility trends in Luzon, Visayas, and Mindanao from 2020 to 2023 (Figure 2). The results highlight how geographic, demographic, and socioeconomic factors shaped the varied responses and outcomes of pandemic-related mobility controls.

**Figure 2:**
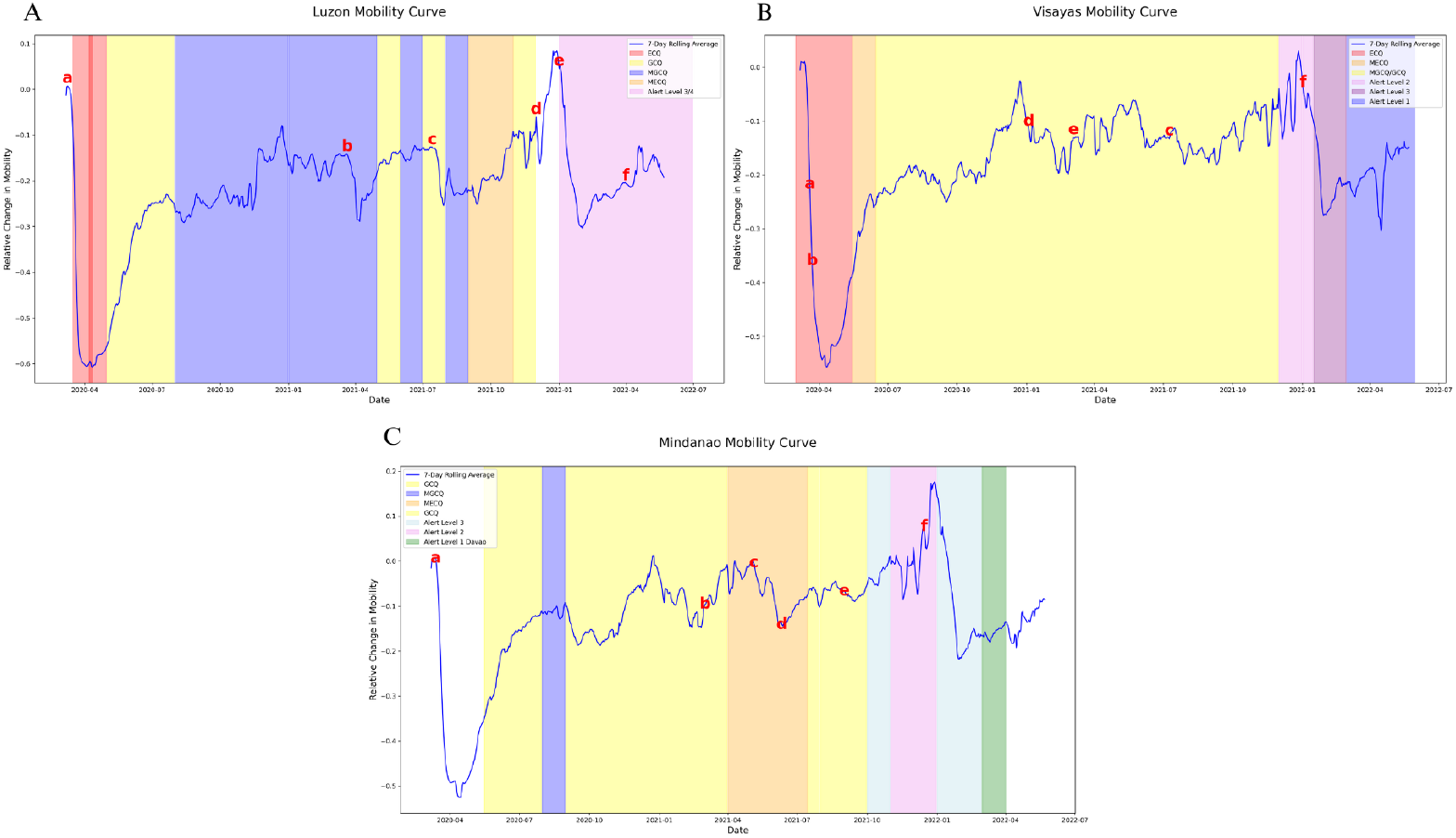
Annotated Mobility Curves in Luzon, Visayas, and Mindanao from 2020–2022. This graph illustrates the relative change in mobility across the three major islands in the Philippines using a 7-day rolling average, overlaid with different quarantine classifications and alert levels during the COVID-19 pandemic. Key points (a–f) mark significant inflection points or changes in the trend corresponding to policy shifts or outbreak waves. **A**. Luzon Mobility Curve. **B**. Visayas Mobility Curve. **C**. Mindanao Mobility Curve.

In Luzon (Figure 2A), a steep and immediate decline in mobility was recorded following the implementation of the Enhanced Community Quarantine (ECQ) in March 2020 (point a), representing the sharpest drop among the three island groups. This rapid contraction reflects the strictness of the government’s initial containment efforts in Luzon, which is home to Metro Manila and serves as the country’s most densely populated and economically significant region (Talabis et al. 2021). While effective in slowing early viral transmission, the mobility reduction also highlighted the difficult balance between protecting public health and sustaining economic activity, especially in urban areas where restrictions had far-reaching effects on livelihoods and services.

Mobility in Luzon recovered unevenly during the shift to General and Modified General Community Quarantine (GCQ and MGCQ) (points b to d). However, the region remained highly sensitive to COVID-19 case surges and corresponding lockdowns. A brief uptick in movement in late 2021 (point e) coincided with the holiday season but was quickly reversed by the arrival of the Omicron variant. By 2022, under the Alert Level System, Luzon’s mobility stabilized at a moderately reduced level (point f). This reflected a mix of behavioral adaptation, growing vaccination coverage, and pandemic fatigue that shaped movement patterns even in the absence of strict enforcement.

Compared to Luzon, the Visayas (Figure 2B) and Mindanao (Figure 2C) displayed more gradual and regionally distinct mobility trends. The Visayas experienced a less abrupt initial drop in movement, followed by a slower but steadier recovery under prolonged GCQ and MGCQ classifications. Although cities like Cebu experienced significant outbreaks, population movement across the region remained relatively consistent. A modest increase in late 2021 (point f) was similarly followed by a drop during the Omicron wave, reflecting similar seasonal and health-related dynamics.

Mindanao demonstrated the most resilient mobility recovery of the three regions. After an initial dip in early 2020 (point a), mobility rebounded more quickly and even rose above baseline levels by mid-2021 (point c). This trend likely reflects the island’s more distributed population, the earlier reopening of key cities such as Davao, and an economy that depends more heavily on agriculture and local trade. Although the introduction of the Alert Level System later curbed mobility to some extent (point f), the impact was less severe than in Luzon.

These regional patterns highlight how mobility responses to the pandemic were shaped not only by national policy but also by local contexts. The move from blanket lockdowns to a more localized, risk-based Alert Level approach signaled a shift in governance strategy. However, differences in enforcement capacity, infrastructure, digital access, and socioeconomic resilience meant that populations across the archipelago experienced and responded to these restrictions in very different ways.

### Evolutionary Dynamics of SARS-CoV-2 Across LuzViMin

To explore the regional evolution of SARS-CoV-2 across the Philippines, we examined phylogenetic trees constructed from genomic sequences collected in Luzon, Visayas, and Mindanao (Figures 3A– 3C). The resulting patterns reveal how viral diversification and transmission dynamics were shaped not only by biological processes, but also by geographic, demographic, and behavioral contexts. Luzon (Figure 3A) displayed the most complex and densely branched phylogenetic structure, with a wide distribution of globally recognized variants, including Alpha (20I), Delta (21A/21J), and Omicron (21K/21L). High sequence density and color dispersion across regions like CALABARZON, Central Luzon, and the Cordillera Administrative Region point to frequent introductions and sustained intra-island transmission. This complexity reflects the status of Luzon Island as the most urbanized and globally connected region in the country, with high mobility volatility (Figure 3A) and consistently elevated case and mortality rates (Figure 1). Notably, periods of rising movement, such as between points d and e in the Luzon mobility curve, preceded major variant surges, suggesting a temporal linkage between increased mobility and variant emergence or amplification.

**Figure 3:**
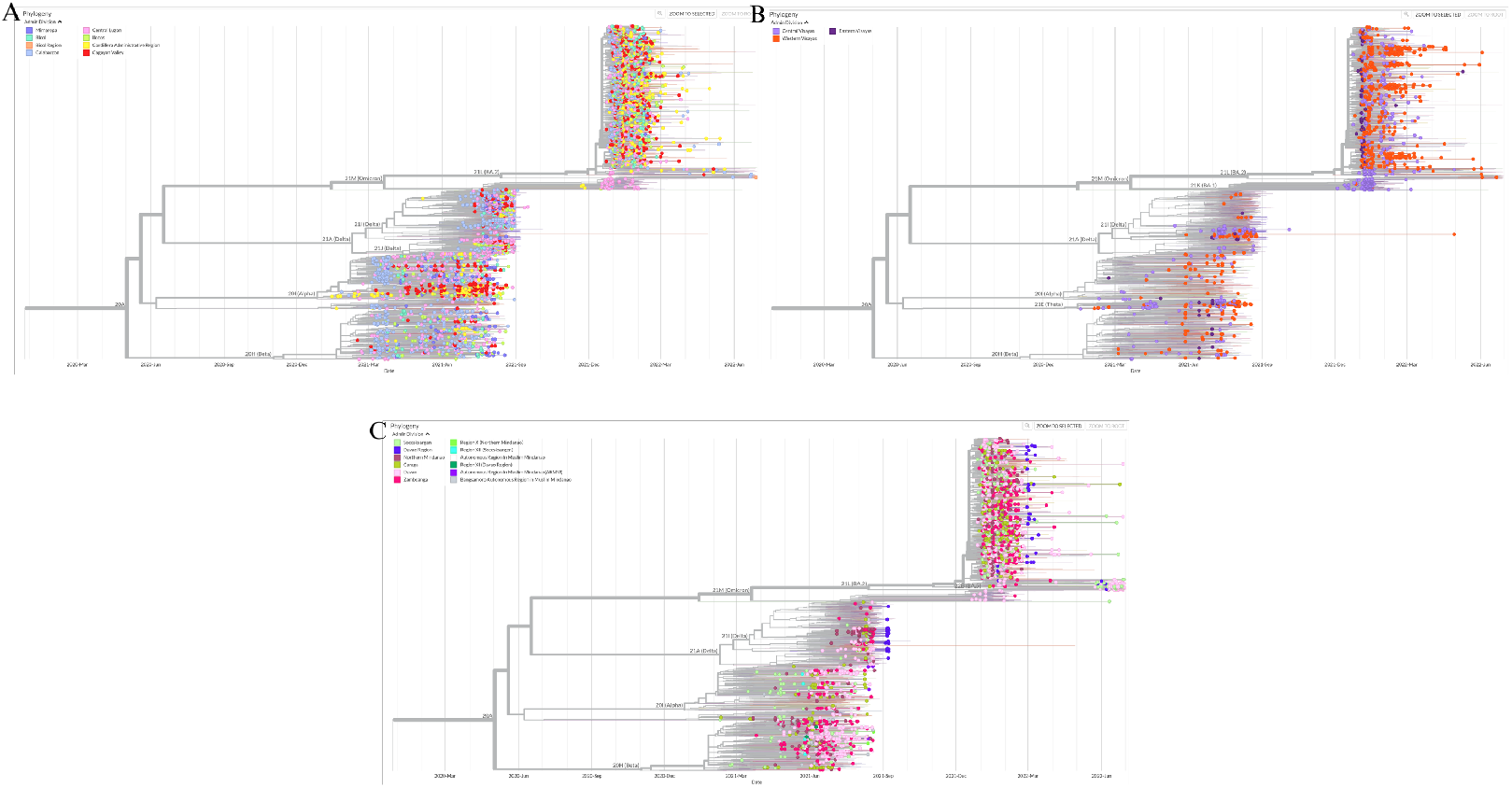
Phylogenetic Trees of SARS-CoV-2 Sequences in Luzon, Visayas, and Mindanao (2020– 2022). The trees are annotated by variant lineage and administrative division. **A**. Luzon Phylogenetic Tree. **B**. Visayas Phylogenetic Tree. **C**. Mindanao Phylogenetic Tree.

In the Visayas (Figure 3B), phylogenetic diversity was notably more limited, with sequences largely clustering around the Delta and Omicron variants. The tree structure reveals fewer and more consolidated branches, implying a smaller number of introduction events and more contained local transmission. This aligns with steadier mobility trends and comparatively flatter epidemic curves in the Visayas region (Figures 1 and 2B). While cities like Cebu served as important regional centers, overall mobility remained more stable following the initial ECQ, and movement spikes were largely isolated to late 2021 (Figure 2B, point f). As such, the evolutionary landscape in the Visayas appears shaped by episodic introductions rather than continuous transmission, and the more subdued branching suggests effective containment of outbreaks before extensive viral diversification could occur. These findings point to a region where spatial fragmentation and decentralized settlement patterns likely served as buffers against widespread viral evolution, by limiting sustained transmission chains and reducing the frequency of variant introductions. Similar dynamics have been observed globally, where rurality, lower population density, and geographic isolation were associated with delayed viral spread and reduced lineage diversity (Dellicour et al. 2021).

Mindanao (Figure 3C) exhibited the most localized and least diverse phylogenetic landscape among the three island groups. Sequences were sparse and primarily derived from a limited set of regions such as Northern Mindanao, Davao, and BARMM, forming compact clusters with minimal branching from major variant lineages. This localized genomic footprint corresponds with the consistently low case and death rates in the Mindanao region, as well as its relatively stable mobility patterns (Figure 3C). The limited inter-island travel, lower urban density, and slower pace of mobility transitions may have reduced opportunities for both variant introduction and sustained community spread. Moreover, earlier relaxation in selected areas like Davao under Alert Level 1 did not appear to substantially alter the evolutionary profile, possibly due to delayed seeding or effective localized control. Taken together, the phylogenetic data reflect a regionally stratified pandemic experience, where viral evolution was most accelerated in areas of high mobility and population density, and markedly constrained in regions with more dispersed populations and lower connectivity.

## CONCLUSION

The spatiotemporal dynamics of COVID-19 in the Philippines reveal a complex interplay between viral evolution, population mobility, and regional demographics. Luzon consistently bore the greatest burden in terms of cases, deaths, and variant diversity, reflecting its dense population, economic centrality, and mobility volatility. In contrast, Visayas and Mindanao exhibited more localized and constrained transmission patterns, with fewer variant introductions and flatter epidemic curves. Mobility data contextualize these trends, highlighting how shifts in movement, driven by quarantine policies and socio-economic structures, shaped the trajectory of the pandemic across island groups. These findings underscore the need for region-specific public health strategies that account for geographic, behavioral, and infrastructural differences during pandemic response and preparedness.

## Data Availability

All data are available in the manuscript.

## ACKNOWLEDGMENTS

Our laboratory is supported by a grant-in-aid from the Philippine Council for Health Research and Development (PCHRD) of the Department of Science and Technology (DOST) of the Philippines.

## CONTRIBUTIONS OF INDIVIDUAL AUTHORS

JKD Aligato, JAB Limbo, GIC Lim, AV Madrigal, and LMS Jingco spearheaded the development of the pipeline used in the study to analyze all three major parameters. They were also responsible for finalizing the models and generating the results of the study. JAB Limbo and JAH Manzano wrote and developed the first draft of the manuscript. RC Cipriano, KPE Cabildo, EMA Allosa, AFD Manrique, DJH Dungo, and ADR Recio contributed to the data collection and curation phases as spearheaded by GIC Lim and AV Madrigal. JAB Limbo, JAH Manzano and N Austriaco revised the manuscript. JAH Manzano and N Austriaco conceptualized, supervised, and oversaw all parts of the conduct of this study. All authors approved the final version of the manuscript.

